# The Effects of Progressive Resistance Strength Training on Pain, Mobility and Activities of Daily Living among Knee Osteoarthritis Patients: A randomized controlled trial

**DOI:** 10.1101/2023.11.20.23298606

**Authors:** Muhammad Tariq Rafiq, Mohamad Shariff A Hamid, Eliza Hafiz, Farid Ahmad Chaudhary, Muhammad Irfan Khan

## Abstract

**Objective:** The objective of this study was to investigate the effects of progressive resistance strength training of lower limb rehabilitation protocol (LLRP) on pain, activities of daily living (ADL) and mobility among knee OA patients who are overweight or obese.

**Materials and Methods:** Fifty-six overweight or obese knee OA patients were included and randomly assigned to a Rehabilitation Protocol Group (RPG) or Control Group (CG). The patients in the RPG performed the progressive resistance strength training of LLRP and followed the instructions of daily care (IDC) for duration of twelve weeks at home. The patients in the CG followed the IDC (conventional treatment) only. Outcome measures were assessed at baseline and after the interventions in both groups by comparing the Western Ontario and McMaster Universities Osteoarthritis Index (WOMAC) score for pain, Timed Up and Go (TUG) test score for mobility and Katz Index of Independence scores for ADL.

**Results:** The patients in the RPG reported significant improvements in WOMAC score for pain (p = 0.001), Katz Index of Independence scores for ADL (p = 0.003) and TUG test score for mobility (p = 0.004). The patients in the CG also reported significant improvements in WOMAC score for pain (p = 0.002) and ADL (p = 0.052), but not in mobility score (p = 0.065). The improvement in the pain and ADL scores was greater in the patients of RPG than the CG with p-value of 0.001 and 0.000 respectively.

**Conclusion:** The progressive resistance strength training of LLRP is effective in terms of reducing pain, improving mobility and ADL among knee OA patients who are overweight or obese.

## INTRODUCTION

In the United States, doctor-diagnosed arthritis is a common chronic condition [1]. A published article pointed out that osteoarthritis (OA) is associated with the joint wear and tear as well as inflammation of the synovial membrane [2]. The most commonly effected joint of lower limb is the knee joint and the exercise intervention is the non-surgical treatment of knee OA joint [3]. Knee OA is a common joint disease, which causes pain and loss of function in elderly persons [4]. A published study explained that the global age-standardized prevalence of knee OA estimated to be 3.8% and it is going to double from the year 2000 to 2020 due to increasing of age population and obesity [5]. Obesity is a significant as well as rapidly increasing global health issue. More than 39% and 13% of adults were considered overweight (body mass index > 25 kg/m2) and obese (body mass index > 30 kg/m2) respectively in 2014, and the prevalence of overweight and obesity has doubled globally since 1980 [6]. A recent published research study concluded that the excess of weight and adiposity had a negative impact in increasing pain perception of patients with OA [7]. .

In knee OA all tissues surrounding the knee joint are being affected with most dominantly the cartilage and subchondral bone of the knee joint. Muscles weakness, narrowing of joint space [8–10], inflammation, osteophyte formation, articular cartilage destruction [8, 10], are the anatomical changes around the knee joint. The research articles pointed out that knee OA patients have 20 to 40 % weaker relative strength of quadriceps muscles compared with control subjects [11, 12]. The biomechanical mal-alignment and weakness of muscles can put abnormal patterns of load stresses inside the knee joint. During walking 70-80% of knee joint loading takes place in the medial compartment of the knee compared to the lateral compartment and if there is mal-alignment and cartilage destruction, then the knee joint loading can increase to 100% in the medial compartment of the knee [13].

The Ottawa Panel found the evidence to support the use of therapeutic exercises, especially strengthening exercises and general physical activity, combined with manual therapy or separately for the improvement of pain for the OA patients [14]. The above study provides the strong evidence for the use of strengthening exercises for the OA patients. In the current study the researcher with the help of expertise prepared a lower limb rehabilitation protocol (LLRP) to strengthen the muscles of lower limbs among knee OA patients who are overweight or obese. A published study provided a strong suggestion of moderate strengthening type exercise for the overweight knee OA patients and they may be able to receive another moderate strengthening type exercise stimulus after only forty-eight hours [15]. A study explained that the progressive resistance strength training increases load gradually over the training course to strengthen the major muscle groups and has been recommended to prevent or reduce late-life disability for older adults [16]. Therefore, in the current study the researcher progressed from low intensity to high intensity exercise gradually by explaining the intensity, frequency and duration of the LLRP in order to reduce pain, improve mobility and activities of daily living (ADLs). Home-based exercise intervention may be effective for relieving Knee OA symptoms [17]. The patients in the current study were instructed to follow their interventions at home. In the current study the researcher applied the implementation of training sessions of progressive resistance strength training of LLRP that are non-weight bearing strengthening exercises in sitting or lying positions and there is minimal mechanic pressure at the knee. Therefore, it is the greatest contribution to the body of knowledge internationally. The purpose of this study was to investigate the effects of progressive resistance strength training of LLRP on pain, mobility and ADLs among knee OA patients who are overweight or obese.

## MATERIALS AND METHODS

### Design and setting

The study was a single blinded randomized controlled trial. The study was conducted in the Teaching Bay of Rehmatul-Lil-Alameen Postgraduate Institute of Cardiology (RAIC), Punjab Employees Social Security Institution (PESSI), Lahore, Pakistan. The study was approved by the reginal Ethical Committee with approval No: RAIC PESSI/Estt/2020/22 and the trial was registered in the Chinese Clinical Trial Registry with registration number: ChiCTR2000034570 on date 10-07-2020. Pre-defined questionnaire of inclusion and exclusion criteria was used for screening of the patients. Patients were recruited from the urban area of Punjab Lahore Pakistan. The experimental procedures, risks and benefits associated with the study were explained to all patients prior to providing written informed consent. Written informed consent was obtained prior to inclusion.

### Study patients

Forty-six overweight or obese knee OA patients completed the study. The inclusion criteria were as follows: overweight (BMI ≥ 25kg/m^2^) or obese (BMI > 30kg/m^2^) [18] knee OA patients; aged between 45-60 years; residing in Urban community of Lahore; history of knee pain for more than 3-months and diagnosed with knee OA of grade 2 or 3 according to Kellgren and Lawrence grading scale [19]. The exclusion criteria were as follows: diagnosed with rheumatoid arthritis, system lupus erythematosus, flat feet or spinal deformities; history of metabolic, hormonal, orthopaedic or cardiovascular disease; previous surgery of knee/s; injections of knee/s for the last six months or unwilling to participate.

### Sample size

Sample size estimation was performed using the G* Power 3.1.3 software. By assuming the medium effect size f = 0.70, setting α = 0.05, power (1-B) = 0.80, number of groups = 2, number of measurements = 2, the total sample size estimated was 33 patients. After considering the apprehensions of drop out or research mortality, the sample size of 56 patients for two groups was decided.

### Patient’s recruitment and randomization

The researcher recruited the patients by active recruitment strategies such as Urban political and welfare organizations via word of mouth by the convenience sampling technique. The list of patients with knee OA in the studied area was obtained from the welfare organizations by explaining the benefits of study participation. Two study coordinators prepared the list of potential patients of knee OA in the recruitment area. After obtaining the list of potential patients of knee OA, the researcher arranged a meeting with the knee OA patients by calling them on the phone. The meeting was held at the Teaching Bay of RAIC, PESSI, Lahore, Pakistan, in the presence of a medical specialist. Patients were screened for eligibility to participate in the study. Only patients fulfilling the inclusion and exclusion criteria of study were invited to participate in this study. After completing the screening of Knee OA patients, the researcher allocated the 56 selected patients into two groups, namely, Rehabilitation Protocol Group (RPG) and Control Group (CG) by a computer-generated random number. Each group consisted of 28 patients.

### Blinding and Allocation

The principle investigator was not blinded in the study. The participants receiving the intervention were kept blinded by simply not informing them of their treatment allocation. The coordinators collecting data were independent individuals from trials and were unaware of the group allocation. There were different coordinators at baseline and after the interventions evaluations. Individuals performing the statistical analysis were kept blinded by labelling the groups with non-identifying terms (such as X and Y).

### Research procedures

#### Research procedure of rehabilitation protocol group (RPG)

The researcher taught the progressive resistance strength training of LLRP and IDC to the patients of RPG for duration of twelve-weeks. Patients were advised to continue performing the progressive resistance strength training of LLRP 3-times a week for twelve-weeks at home. These trainings included the strengthening exercises for the lower limbs in non-weight bearing sitting or lying positions. Each training session started with ten minutes warm up, forty-five to sixty minutes of lower limbs progressive resistance training, and a ten minutes cool down at the end of training protocol.

A cool-down period was essential after a training session and should last approximately 5 to ten minutes [20, 21]. When the static stretching was used as part of a warm-up immediately prior to exercise, then it causes harm to muscle strength [22]. The patients in the RPG performed the progressive resistance strength training of LLRP and followed the IDC at home for duration of twelve-weeks. The IDC are explained elsewhere [23].

The sequence of the training programme started with a ten minutes’ warm-up with whole body range of motion (ROM) and dynamic stretching exercises (Table 1). The patients performed ten repetitions of ROM of each muscle group and 5 repetitions of dynamic stretching of each muscle group as a part of warm-up. After the warm-up the participants performed the strengthening exercises of LLRP for stipulated weeks as stated in Table 2. The patients followed the IDC, which are explained earlier [23]. After completing the strengthening exercises, the patients performed the ten minutes cool-down with whole body ROM and static stretching exercises (Table 1). The patients performed ten repetitions of ROM of each muscle group and 3 repetitions of static stretching of each muscle group as a part of cool-down.

**Table 1.**
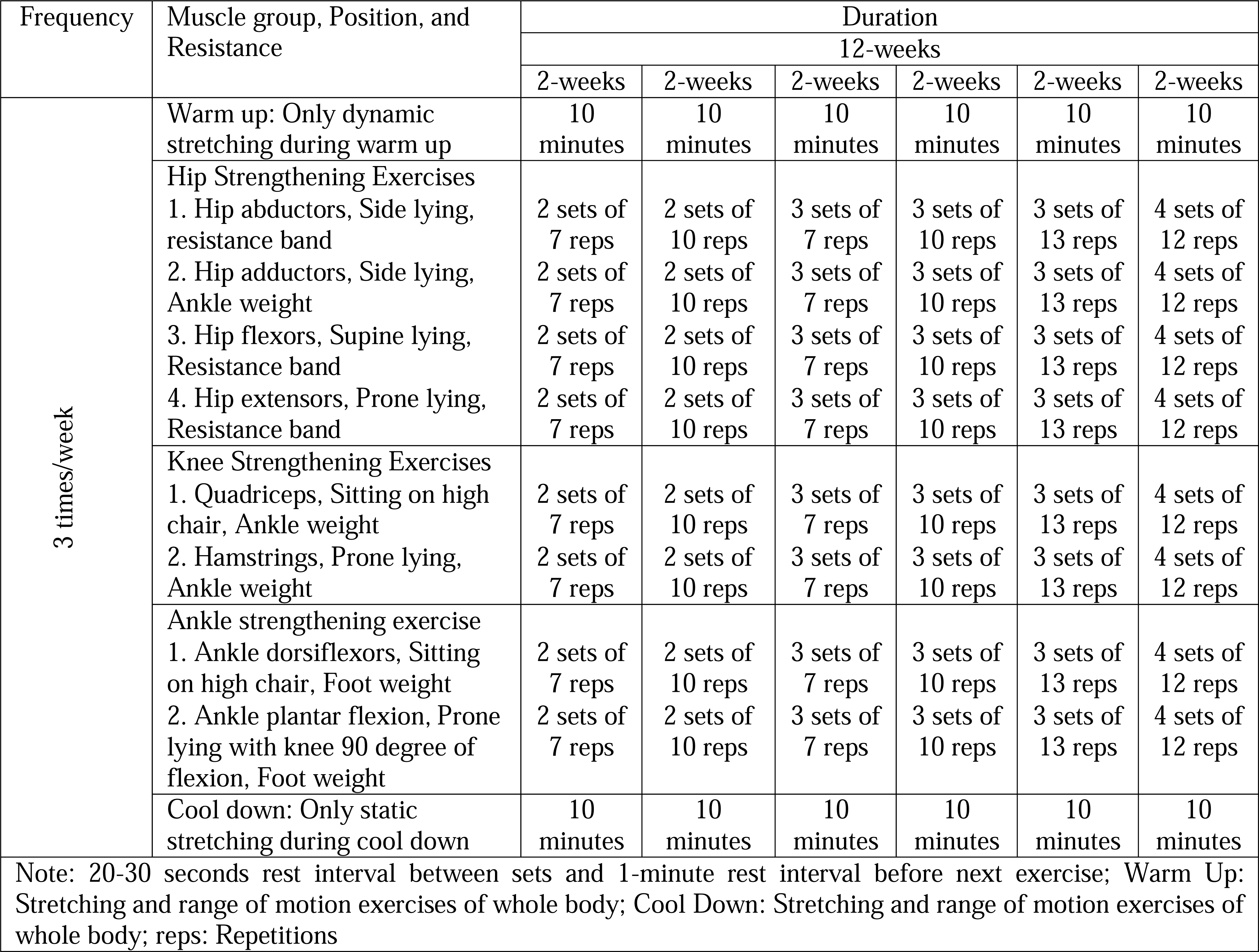
Lower Limb Rehabilitation Protocol (LLRP)

**Table 2.** Baseline socio-demographic characteristics and clinical outcome measures of patients: mean (SD), median (IQR) or No (%)

| **Socio-demographic characteristics** | **RPG**<br>(n = 23) | **CG**<br>(n = 23) | **P-value** |
| --- | --- | --- | --- |
| **Age**, mean (SD), y | 53.78 (5.38) | 54.56 (4.91) | 0.609 |
| **Gender** (M/F) | 9/14 | 8/15 | 1.000 |
| **Educational Status**, No. (%) |  |  |  |
| Matriculation | 8(34.8) | 1 (4.3) | 0.293 |
| Intermediate | 9 (39.1) | 15 (65.2) |  |
| Bachelor | 3(13.0) | 4 (17.4) |  |
| Master | 3(13.0) | 3 (13.0) |  |
| **Marital status**, No. (%) |  |  |  |
| Single | 3 (13.0) | 0 | 0.865 |
| Married | 18 (78.3) | 22 (95.7) |  |
| Divorced | 2 (8.7) | 1 (4.3) |  |
| **Socioeconomic status (According to Pakistan Rupees),**<br>No. (%) |  |  |  |
| Lower | 17 (73.9) | 16 (69.6) | 0.649 |
| Middle | 4 (17.4) | 4 (17.4) |  |
| Upper | 2 (8.7) | 3 (13.0) |  |
| **WOMAC Pain score (0-20)**, median (IQR) | 8.00 (3.00) | 10.00 (3.00) | 0.547 |
| **Mobility (TUG test score in seconds)**, median (IQR) | 10.00 (5.00) | 9.30 (4.00) | 0.535 |
| **Katz index of independence in ADL score**, median<br>(IQR) | 4.00 (2.00) | 4.00 (2.00) | 0.945 |

Abbreviation: RPG = Rehabilitation Protocol Group; CG = Control Group; WOMAC = Western Ontario and McMaster Universities Osteoarthritis Index; SD = Standard Deviation; IQR = Interquartile Range; TUG = Timed Up and Go; ADL = Activities of Daily Living; N = Number

#### Research procedure of control group (CG)

The patients in the CG were not involved in the rehabilitation protocols, but these patients only followed the IDC for duration of twelve-weeks. Two language experts also translated the IDC into Urdu language as the patients’ preference of the Urdu translation for better understanding based on a recent pilot study [23]. All patients were also given a diary and asked to record the attendance of completion their interventions based on leaflets.

#### Outcome measures

Patients’ socio-demographics, pain, mobility and ADL were taken at baseline before randomization. At the end of twelve-weeks, the identical measurements of socio-demographics, pain, mobility and ADL were repeated. The socio-demographic questionnaire covered age, gender, educational status, marital status and socioeconomic status. Outcome measures were categorized into the primary and secondary outcome measures.

#### Primary Outcome Measures

These were pain and ADL. The pain was assessed using the Western Ontario and McMaster Universities Osteoarthritis Index (WOMAC) score for pain. The WOMAC score ranges from 0 to 4 on a Likert-type scale, the higher the score, the worse the pain. The questions asked to evaluate the level of pain scores in the pain part of WOMAC score comprised; walking on a flat surface, going up or down stairs, at night while in bed, sitting or lying and standing upright as well as level of pain in the past 48 hours in the right and left knee [24].

The Katz Index of Independence in ADL was used for the measurement of ADL. In Katz ADL 6 functions were assessed such as bathing, dressing, toileting, transferring, continence, feeding. One point was used as independence with subheadings no supervision, direction or personal assistance. Zero point was used as dependence with subheadings supervision, direction, personal assistant or total care. In Katz index of independence in ADL a score of 2 or less indicates severe functional impairment, 4 indicates moderate impairment and score of 6 indicates full functional independence in ADL [25].

#### Secondary outcome measures

The secondary outcome measure was the mobility. Time Up and Go (TUG) test was applied for the measurement of mobility. The patients were observed to note their mobility time while they rose from an arm chair, walked 3 meters, turned, walked back, and sat down again. During the process standardized instructions for testing as described in the literature were followed [26].

#### Statistical procedures

Statistical Package for Social Sciences (SPSS), version 22, Chicago, IL, was used to analyze the data. Descriptive statistics were expressed as the percentage (%), mean, standard deviation (SD), median and interquartile range (IQR). Inferential statistics was used for all the quantitative measures. Prior to data analysis, Shapiro-Wilk test was used for all variables to check the normality of data. The scores were not normally distributed; therefore, the Wilcoxon Signed Ranked Test was used to analyze the data within groups. The Mann Whitney U - test was used to evaluate the differences between the groups. Value of P ≤ 0.05 was considered to be statistically significant.

## RESULTS

Figure 1 shows a flow chart of patients’ participation in this study. A total of seventy-five overweight and obese knee OA patients were initially inducted after assessing for eligibility by a pre-defined inclusion and exclusion criteria. Among them, nineteen patients (5 due to normal weight, 2 with a past history of cardiovascular disease, 3 due to injections of knee, 1 due to spinal deformities, 4 due to Kellgren and Lawrence grades, 2 due to knee surgery and 2 being unwilling to participate) were excluded; the remaining fifty-six patients were randomized equally into 2 groups. Of the twenty-eight patients allocated to the RPG, 5 did not continue their intervention because they were unable to continue: 3 were sick and remaining 2 were at outstation. Likewise, 5 of the twenty-eight patients allocated to the CG did not continue IDC because 3 got the urine problem and remaining 2 were unwilling to continue due to domestic problems. We could not obtain the post-intervention outcomes for these ten withdrew patients. A resultant total of forty-six patients (twenty-three in the RPG and twenty-three in the CG) completed the study.

**Figure 1.**
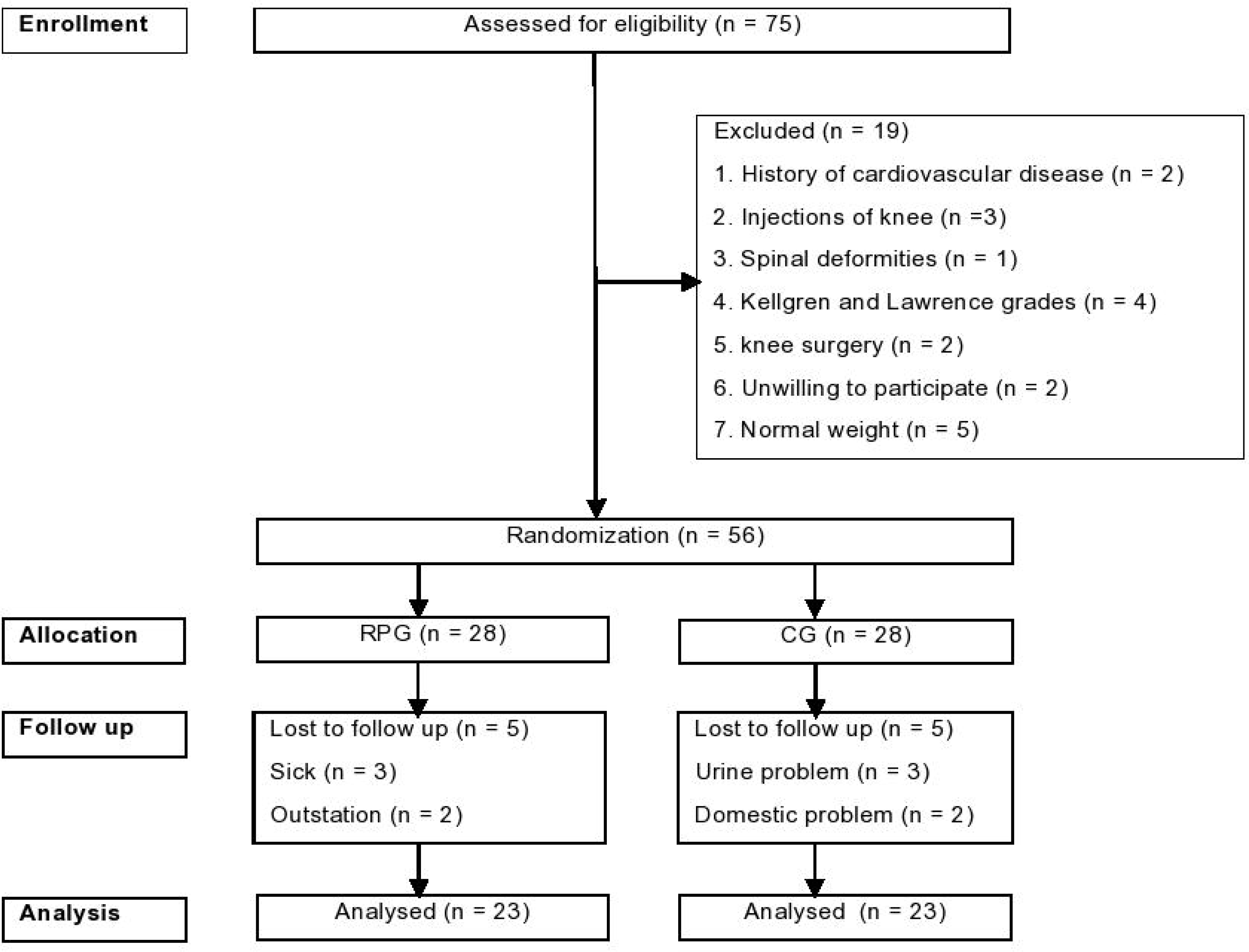

Baseline socio-demographic characteristics and clinical outcome measures are presented in Table 2. No significant differences were observed in baseline socio- demographic characteristics. Similarly, baseline clinical outcome measures of pain, mobility and ADL between the two groups showed no significant differences (Table 2).

After 12 weeks of training sessions of strengthening exercises of LLRP, the patients in the RPG reported significant improvements in the WOMAC pain score (p = 0.001), TUG test score (p = 0.004) and Kats index of independence in ADL scores (p = 0.003) (Table 3).

**Table 3.** Comparison of the rehabilitation protocol group (RPG) and control group (CG) variables at 12 weeks.

| Variables | RPG (n = 23) |  |  |  | CG (n = 23) |  |  |  |
| --- | --- | --- | --- | --- | --- | --- | --- | --- |
|  | Median (IQR) | Median of the difference (95 % CI) | Z-test | P-value | Median (IQR) | Median of the difference (95 % CI) | Z-test | P-value |
| WOMAC Pain score (0 to 20) |  |  |  |  |  |  |  |  |
| Baseline | 8.00 (3.00) | -2.00 (-3.00 to -1.50) | -3.46 | 0.001 | 10.00 (3.00) | -0.50 (-1.00 to 0.00) | -3.06 | 0.002 |
| After the intervention | 11.00 (4.00) |  |  |  | 10.00 (3.00) |  |  |  |
| Mobility (TUG Test in seconds) score |  |  |  |  |  |  |  |  |
| Baseline | 10.00 (5.00) | -1.15 (-2.20 to -0.45) | -2.89 | 0.004 | 9.30 (4.00) | -0.05 (-0.15 to 0.00) | -1.84 | 0.067 |
| After the intervention | 9.00 (5.00) |  |  |  | 9.05 (4.00) |  |  |  |
| Katz index of independence in ADLs score |  |  |  |  |  |  |  |  |
| Baseline | 4.00 (2.00) | 1.00 (0.50 to 1.50) | -2.96 | 0.003 | 4.00 (2.00) | -0.50 (-0.00 to 1.00) | -1.94 | 0.051 |
| After the intervention | 5.00 (1.00) |  |  |  | 4.00 (1.00) |  |  |  |

Abbreviation: RPG = Rehabilitation Protocol Group; CG = Control Group; WOMAC = Western Ontario and McMaster Universities Osteoarthritis Index; CI = Confidence Interval; IQR = Interquartile Range; TUG = Timed Up and Go; ADL = Activities of Daily Living

Similarly, a significant improvement was also reported in the WOMAC pain score (p = 0.002) and Kats index of ADL scores (p = 0.052) by the CG. By contrast, no significant improvement was reported by TUG test score (p = 0.065) in the CG (Table 3).

Comparison of outputs between the 2 groups, RPG group revealed significant improvement in the pain and ADL scores than the CG, the ample cause of which was the p values for the pain (p = 0.001) and ADL (p = 0.000) scores. However, the RPG did not report significant in the mobility score than the CG because the P value was 0.155 (Table 4).

**Table 4.** Comparison between the RPG and CG.

| **Outcomes** | **RPG** |  | **CG** |  | **P-value** |
| --- | --- | --- | --- | --- | --- |
|  | Baseline | After the intervention | Baseline | After the intervention |  |
| **WOMAC Pain (0-20) score, median (IQR)** | 8.00 (3.00) | 11.00 (4.00) | 10.00 (3.00) | 10.00 (3.00) | 0.001 |
| **Mobility (TUG Test in seconds) score, median (IQR)** | 10.00 (5.00) | 9.00 (5.00) | 9.30 (4.00) | 9.05 (4.00) | 0.155 |
| **Katz index of independence in ADLs score, median (IQR)** | 4.00 (2.00) | 5.00 (1.00) | 4.00 (2.00) | 4.00 (1.00) | 0.000 |

Abbreviation: RPG = Rehabilitation Protocol Group; CG = Control Group; WOMAC = Western Ontario and McMaster Universities Osteoarthritis Index; IQR = Interquartile Range; TUG = Timed Up and Go; ADL = Activities of Daily Living

## DISCUSSION

A study reported that home exercise programs with or without clinical-based exercises are commonly used in clinical physiotherapy practice among individuals with knee OA [27]. Adequate number of studies to substantiate the importance of home exercise programs may not support this fact. The study under discussion expressed the effectiveness of home rehabilitation protocol that pertained to progressive resistance strength training of lower limbs conducted by a physiotherapist. The results of current study divulged reduction in pain and improvement in ADL in the patients of RPG and CG groups, on the other hand the improvement in mobility could be seen only in the patients of RPG, but not in the CG. The Study under discussion indicate that, inclusive of the progressive resistance strength training of LLRP in non-weight bearing sitting or lying positions could reduce pain, improve mobility and ADL more efficiently than could usual care.

Another reputed study points to the fact that the greater the intensity of exercise, the greater would be the output effects on knee OA in rats [28]. In the current study, the intensity of training sessions of lower limb rehabilitation protocol progressed from low to high intensity gradually as shown in table 1, to study the progressive resultant change due to successive intensity of exercise. A published experimental study recommended the easier exercises in order to perform and continue with low-intensity exercises for knee OA patients [29].

In a systematic review and meta-regression analysis of randomized controlled trial, it was reported that optimal exercise program should focus on improving lower extremity performance or quadriceps muscle strength among knee OA patients [30]. Therefore, the training sessions of progressive resistance strength training of LLRP were used in the current study.

A systematic review that investigated the impact of exercise type and dose in knee OA divulged that exercise sessions were between 1 to 5 training sessions per week with the duration of exercise lasting between 3 weeks to 9 months [30]. In the current study, the training sessions of LLRP were conducted thrice a week continuing for 12 weeks. Two systematic reviews of randomized controlled trials have reported that exercise therapy reduces pain for OA of the knee [30, 31]. In the current study, the score of pain was significantly reduced in both the patients of RPG and CG, but marked reduction in pain score was reported by the patients in the RPG rather than the CG group.

A randomized controlled trial reported that the cumulative of diet plus exercise group improvement in mobility was significantly greater than either the solo diet or solo exercise group [32] as well as greater than that reported by the diet plus exercise group of the arthritis, diet, and activity promotion trial [33]. The current study divulged that the patients in the RPG did not report more significant improvement in the mobility scores than the CG. The rating of Katz ADL is recommended by the observation of the health care professionals [34]. In the current study, a trained health professional recorded the score of Katz ADL. The patients in the RPG reported more significant improvement in ADL than the CG.

This current randomized controlled trial provided further substantiation that the training sessions of progressive resistance strength training of LLRP based on muscle group exercises in non-weight bearing sitting or lying positions proved more effectiveness in reducing pain and improving ADLs than typical rehabilitation.

This study had several limitations. First, the results may not be generalized to all overweight or obese knee OA patients because inducted patients were with second and third degree severity of knee OA according to Kellgren and Lawrence grading scale [19]. No long-term follow-up records were taken. Finally, the comparisons were made on a relatively smaller number of patients in this study. Thus, further research on a larger sample size and long-term follow-up is required to confirm the results of the progressive resistance strength training of LLRP.

### Funding

The authors received no financial support for the research and/or authorship of this article.

### Data Availability

The data used to support the findings of the study are available from the corresponding author upon request.

## Supporting information

www.consort-statement.org

## REFERENCES

1. Barbour KE, Helmick CG, Boring M, Zhang X, Lu H, Holt JB. Prevalence of doctor-diagnosed arthritis at state and county levels—United States, 2014. Morb Mortal Wkly Rep. 2016;65(19):489-94. doi: 10.15585/mmwr.mm6519a2

2. Helmark IC, Mikkelsen UR, Børglum J, Rothe A, Petersen MC, Andersen O, Langberg H, Kjaer M. Exercise increases interleukin-10 levels both intraarticularly and peri-synovially in patients with knee osteoarthritis: a randomized controlled trial. Arthritis Res Ther. 2010;12(4): R126. 10.1186/ar3064

3. National Clinical Guideline Centre (UK). Osteoarthritis: Care and Management in Adults. London: National Institute for Health and Care Excellence (UK); 2014. PMID: 25340227

4. McAlindon TE, Bannuru R, Sullivan MC, Arden NK, Berenbaum F, Bierma-Zeinstra SM, Hawker GA, Henrotin Y, Hunter DJ, Kawaguchi H, Kwoh K. OARSI guidelines for the non-surgical management of knee osteoarthritis. Osteoarthr Cartilage. 2014;22(3):363–88. 10.1016/j.joca.2014.01.003

5. Cross M, Smith E, Hoy D, Nolte S, Ackerman I, Fransen M, Bridgett L, Williams S, Guillemin F, Hill CL, Laslett LL. The global burden of hip and knee osteoarthritis: estimates from the global burden of disease 2010 study. Ann Rheum Dis. 2014;73(7):1323–30. 10.1136/annrheumdis-2013-204763

6. Finucane MM, Stevens GA, Cowan MJ, Danaei G, Lin JK, Paciorek CJ, Singh GM, Gutierrez HR, Lu Y, Bahalim AN, Farzadfar F. National, regional, and global trends in body-mass index since 1980: systematic analysis of health examination surveys and epidemiological studies with 960 country-years and 9·1 million participants. Lancet. 2011 Feb 12;377(9765):557-67. 10.1016/S0140-6736(10)62037-5

7. Alfieri FM, Silva NC, Battistella LR. Study of the relation between body weight and functional limitations and pain in patients with knee osteoarthritis. Einstein (Sao Paulo). 2017;15(3):307–12. 10.1590/s1679-45082017ao4082

8. Arden N, Nevitt MC. Osteoarthritis: epidemiology. Best Pract Res Clin Rheumatol. 2006;20(1):3–25. 10.1016/j.berh.2005.09.007

9. Pearle AD, Warren RF, Rodeo SA. Basic science of articular cartilage and osteoarthritis. Clin Sports Med. 2005;24(1):1–2. 10.1016/j.csm.2004.08.007

10. Sharma L, Kapoor D, Issa S. Epidemiology of osteoarthritis: an update. Curr Opin Rheumatol. 2006;18(2):147–56. doi: 10.1097/01.bor.0000209426. 84775.f8

11. Jamtvedt G, Dahm KT, Christie A, Moe RH, Haavardsholm E, Holm I, Hagen KB. Physical therapy interventions for patients with osteoarthritis of the knee: an overview of systematic reviews. Phys Ther. 2008;88(1):123–36. 10.2522/ptj.20070043

12. Rutjes AW, Nüesch E, Sterchi R, Kalichman L, Hendriks E, Osiri M, Brosseau L, Reichenbach S, Jüni P. Transcutaneous electrostimulation for osteoarthritis of the knee. Cochrane Database Syst Rev. 2009(4). 10.1002/14651858.CD002823.pub2

13. Schipplein OD, Andriacchi TP. Interaction between active and passive knee stabilizers during level walking. J Orthop Res. 1991;9(1):113–9. 10.1002/jor.1100090114

14. Ottawa Panel Members, Ottawa Methods Group, Brosseau L, Wells GA, Tugwell P, Egan M, Dubouloz CJ, Casimiro L, Robinson VA, Pelland L, McGowan J. Ottawa panel evidence-based clinical practice guidelines for therapeutic exercises and manual therapy in the management of osteoarthritis. Phys Ther. 2005;85(9):907–71. 10.1093/ptj/85.9.907

15. Germanou EI, Chatzinikolaou A, Malliou P, Beneka A, Jamurtas AZ, Bikos C, Tsoukas D, Theodorou A, Katrabasas I, Margonis K, Douroudos I. Oxidative stress and inflammatory responses following an acute bout of isokinetic exercise in obese women with knee osteoarthritis. Knee. 2013;20(6):581–90. 10.1016/j.knee.2012.10.020

16. Seguin R, Nelson ME. The benefits of strength training for older adults. Am J Prev Med. 2003;25(3):141–9. 10.1016/S0749-3797(03)00177-6

17. Chen H, Zheng X, Huang H, Liu C, Wan Q, Shang S. The effects of a home-based exercise intervention on elderly patients with knee osteoarthritis: a quasi-experimental study. BMC Musculoskelet Disord. 2019 Dec;20(1):160. 10.1186/s12891-019-2521-4.

18. 18. World Health Organization EC. Appropriate body-mass index for Asian populations and its implications for policy and intervention strategies. Lancet (London, England). 2004;363(9403):157. 10.1016/S0140-6736(03)15268-3

19. Kellgren JH, Lawrence JS. Radiological assessment of osteo-arthrosis. Ann Rheum Dis. 1957;16(4):494. doi: 10.1136/ard.16.4.494

20. Powers SK, Howley ET. Exercise physiology: Theory and application to fitness and performance. New York, NY: McGraw-Hill; 2007.

21. Prentice WE. The thigh, hip, groin, and pelvis. Arnheim’s principles of athletic training: A competency-based approach. 2003:656–98.

22. McHugh MP, Nesse M. Effect of stretching on strength loss and pain after eccentric exercise. Med Sci Sports Exerc. 2008;40(3):566–73. doi: 10.1249/mss.0b013e31815d2f8c

23. Rafiq MT, A Hamid MS, Hafiz E, Amin S. Rehabilitation protocol with or without mobile health in overweight and obese knee osteoarthritis patients-a pilot study. Balneo Research Journal. 2019;10(4):580–584. 10.12680/balneo.2019.306

24. Alexandre Tda S, Cordeiro RC, Ramos LR. Factors associated to quality of life in active elderly. Rev Saude Publica. 2009;43(4):613–621. doi:10.1590/s0034-89102009005000030

25. Shelkey M, Wallace M. Katz Index of Independence in Activities of Daily Living. J Gerontol Nurs. 1999;25(3):8–9. doi:10.3928/0098-9134-19990301-05

26. Podsiadlo D, Richardson S. The timed “Up & Go”: a test of basic functional mobility for frail elderly persons. J Am Geriatr Soc. 1991;39(2):142–8. 10.1111/j.1532-5415.1991.tb01616.x

27. Maher CG, Sherrington C, Herbert RD, Moseley AM, Elkins 463 M. Reliability of the PEDro Scale for rating quality of randomized controlled trials. Phys Ther. 2003; 83(8): 713–721.

28. Li XD, Sun GF, Zhu WB, Wang YH. Effects of high intensity exhaustive exercise on SOD, MDA, and NO levels in rats with knee osteoarthritis. Genet Mol Res. 2015;14(4):12367-12376. Published 2015 Oct 16. doi: 10.4238/2015.October.16.3

29. Kuru Çolak T, Kavlak B, Aydoğdu O, et al. The effects of therapeutic exercises on pain, muscle strength, functional capacity, balance and hemodynamic parameters in knee osteoarthritis patients: a randomized controlled study of supervised versus home exercises. Rheumatol Int. 2017;37(3):399–407. doi:10.1007/s00296-016-3646-5

30. Juhl C, Christensen R, Roos EM, Zhang W, Lund H. Impact of exercise type and dose on pain and disability in knee osteoarthritis: a systematic review and meta-regression analysis of randomized controlled trials. Arthritis Rheumatol. 2014;66(3):622–636. doi:10.1002/art.38290

31. Fransen M, McConnell S, Harmer AR, Van der Esch M, Simic M, Bennell KL. Exercise for osteoarthritis of the knee: a Cochrane systematic review. Br J Sports Med. 2015;49(24):1554–1557. doi:10.1136/bjsports-2015-095424

32. Messier SP, Mihalko SL, Legault C, et al. Effects of intensive diet and exercise on knee joint loads, inflammation, and clinical outcomes among overweight and obese adults with knee osteoarthritis: the IDEA randomized clinical trial. JAMA. 2013;310(12):1263–1273. doi:10.1001/jama.2013.277669

33. Messier SP, Loeser RF, Miller GD, et al. Exercise and dietary weight loss in overweight and obese older adults with knee osteoarthritis: The Arthritis, Diet, and Activity Promotion Trial. Arthritis Rheum. 2004;50(5):1501–1510. doi:10.1002/art.20256

34. Katz S, Ford AB, Moskowitz RW, Jackson BA, Jaffe MW. Studies of illness in the aged: the index of ADL: a standardized measure of biological and psychosocial function. JAMA. 1963; 185:914–919. doi:10.1001/jama.1963.03060120024016

